# Feasibility to virtually generate T2 fat-saturated breast MRI by convolutional neural networks

**DOI:** 10.1101/2024.06.25.24309404

**Authors:** Andrzej Liebert, Dominique Hadler, Chris Ehring, Hannes Schreiter, Luise Brock, Lorenz A. Kapsner, Jessica Eberle, Ramona Erber, Julius Emons, Frederik B. Laun, Michael Uder, Evelyn Wenkel, Sabine Ohlmeyer, Sebastian Bickelhaupt

## Abstract

**Background:** Breast magnetic resonance imaging (MRI) protocols often include T2-weighted fat-saturated (T2w-FS) sequences, which are vital for tissue characterization but significantly increase scan time.

**Purpose:** This study aims to evaluate whether a 2D-U-Net neural network can generate virtual T2w-FS images from routine multiparametric breast MRI sequences.

**Materials and Methods:** This IRB approved, retrospective study included n=914 breast MRI examinations performed between January 2017 and June 2020. The dataset was divided into training (n=665), validation (n=74), and test sets (n=175). The U-Net was trained on T1-weighted (T1w), diffusion-weighted imaging (DWI), and dynamic contrast-enhanced (DCE) sequences to generate virtual T2w-FS images (VirtuT2). Quantitative metrics and a qualitative multi-reader assessment by two radiologists were used to evaluate the VirtuT2 images.

**Results:** VirtuT2 images demonstrated high structural similarity (SSIM=0.87) and peak signal-to-noise ratio (PSNR=24.90) compared to original T2w-FS images. High level of the frequency error norm (HFNE=0.87) indicates strong blurring presence in the VirtuT2 images, which was also confirmed in qualitative reading. Radiologists correctly identified VirtuT2 images with 92.3% and 94.2% accuracy, respectively. No significant difference in diagnostic image quality (DIQ) was noted for one reader (p=0.21), while the other reported significantly lower DIQ for VirtuT2 (p<=0.001). Moderate inter-reader agreement was observed for edema detection on T2w-FS images (ƙ=0.43), decreasing to fair on VirtuT2 images (ƙ=0.36).

**Conclusion:** The 2D-U-Net can technically generate virtual T2w-FS images with high similarity to real T2w-FS images, though blurring remains a limitation. Further investigation of other architectures and using larger datasets are needed to improve clinical applicability.

**Summary Statement:** Virtual T2-weighted fat-saturated images can be generated from routine breast MRI sequences using convolutional neural networks, showing high structural similarity but with notable blurring, necessitating further refinement for clinical use.

**Key Results:**

1. Images with T2w-FS characteristics can be virtually generated from T1w and DWI images using deep learning
2. Image blurring occurring in the VirtuT2 image limit clinical use for the current moment
3. Further investigation of different architectures and with larger datasets are necessary in the future to improve the VirtuT2 performance.

## INTRODUCTION

Breast magnetic resonance imaging (MRI) commonly utilizes a multiparametric protocol [1]. Such multiparametric protocols typically comprises different imaging sequences: unenhanced T1-weighted (T1w), T2-weighted fat-saturated (T2w-FS), diffusion-weighted imaging (DWI, with at least two b-values) and multiple T1w contrast-enhanced (T1w-CE) sequences that form the dynamic contrast-enhanced (DCE) series.

Among those acquisition sequences, the T2w-FS acquisition provides morphological insight into tissue composition and fluid presence, aiding in lesion characterization and being included in classification schemes such as the Kaiser-Score [1]. However, acquiring the T2w-FS images with sufficient spatial resolution can significantly contribute to the total scanning time, sometimes accounting for up to 20% of the entire breast MRI examination [2; 3]. This influences scanner throughput, which is of relevance especially when aiming to use breast MRI as a supplemental screening modality in breast cancer screening programs [4].

In recent years several publications showed that deep-learning approaches are technically feasible to derive “virtual” contrast-enhanced images from multiparametric unenhanced acquisitions in the breast [5-10]. The aim of these virtual contrast-enhanced (vCE) techniques, in all body regions, is focused on the reduction or even potential elimination of gadolinium-based contrast agents (GBCA) administration [11] in order to reduce the costs, potential side effects [12], and environmental issues[13] arising from it. These vCE methods, show the potential to synthesize additional or missing [14-16] diagnostic information from existing MRI acquisitions, thus potentially streamlining the imaging process.

This feasibility study builds on these recent works by investigating, if a 2D-U-Net architecture might be used to generate images mimicking T2w-fat-satured images from other routine sequences used in breast MRI such as T1w, DWI, and dynamic contrast enhanced sequences (DCE). We evaluate such virtual T2w-mimicking images (called “VirtuT2”) both in a quantitative analysis of image similarity and error metrics, and in a multi-reader qualitative assessment study.

## MATERIALS AND METHODS

### Patient cohort and acquisition protocol

This retrospective study has been approved by the ethics committee of Friedrich-Alexander University Erlangen-Nürnberg waiving the need for informed consent. All included breast MRI examinations were clinically indicated routine examinations, performed between January 2017 and June 2020 (age 52±12 years) at the Institute of Radiology, University Clinic Erlangen. The clinical indications included preoperative exclusion of multifocal disease, screening in women with positive family history of breast cancer, exclusion of recurrent breast cancer, clarification of unclear findings in mammography, ultrasound or due to clinical complaints.

The examinations were performed using one of two routine 3T MRI scanners (Magentom Skyra fit or Magnetom Vida, Siemens Healthineers, Erlangen, Germany). Routine multiparametric breast MRI protocol including unenhanced T1w-DIXON, T2w-FS, and multi-b-value DWI (b-values=50, 750, and 1500 s/mm^2^) acquisitions together with five T1-weigthed DCE acquisitions performed in 60 seconds interval after the intravenous administration of GBCA. Due to artifacts which are caused by the silicone in DWI acquisitions, the presence of breast implants was defined as an exclusion criterion. Detailed MRI acquisition parameters are available in Table 1.

**Table 1:** MRI Protocol.

| Sequence | Sequence Type | Matrix Size | FoV (mm×mm) | Slice thickness (mm) | TR (ms) | TE (ms) | IR (ms) | FA (°) | No. of Averages | Fat Saturation |
| --- | --- | --- | --- | --- | --- | --- | --- | --- | --- | --- |
| T1w* | 3D-GRE | 448×448×112–128 | 360×360–430×430 | 1.5–1.8 | 5.97 | 2.46 | - | 10 | 1 | None |
| T1w-FS | 3D-GRE | 448×448×112–128 | 360×360–430×430 | 1.5–1.8 | 5.97 | 2.46 | - | 10 | 1 | DIXON water-phase |
| T2w-FS | 2D-SE | 448×448×34–49 | 340×340–430×430 | 4 | 3570–5020 | 60, 70 | 230 | 108 | 2 | STIR |
| DWI** | 2D-IR-DWI-EPI | 256×160–200×34–49 | 350×219–430×269 | 4 | 6290–9660 | 66, 70 | 220, 250 |  | 3/8/20 or 3/8/15*** | STIR |

FoV=field of view, TR=repetition time, TE=echo time, IR=inversion recovery time, DWI=diffusion-weighted imaging, GRE=gradient echo, SE=spin echo, EPI=echo-planar imaging, GBCA=gadolinium-based contrast agent, STIR=short TI inversion recovery
\*Acquired before and during the five timepoints after intravenous GBCA injection (gadobutrol; Bayer, Leverkusen, Germany; 0.1 mmol/kg/body weight, injection speed=2 mL/s) \*\*DWI scan was acquired using three b-values: 50, 750, and 1500 s/mm<sup>2</sup> \*\*\*No. of averages for DWI indicates the number of averages for each b-value. The first set of values is for the acquisition performed on Skyra-Fit and the second set for that performed on MAGNETOM VIDA.

Detailed definition of the final cohort (n=914) based on the inclusion and exclusion criteria is presented in Figure 1. The dataset was randomly divided at the examination level into a training (n=665), validation (n=74) and independent test set (n=175). During the split it was ensured that no data-leakage on the patient level occurred between the datasets. Optimization of the training hyperparameters was performed based on the performance on the validation set. The independent test set was kept separately until the fixed model was deployed for evaluation of this study.

**Figure 1:**
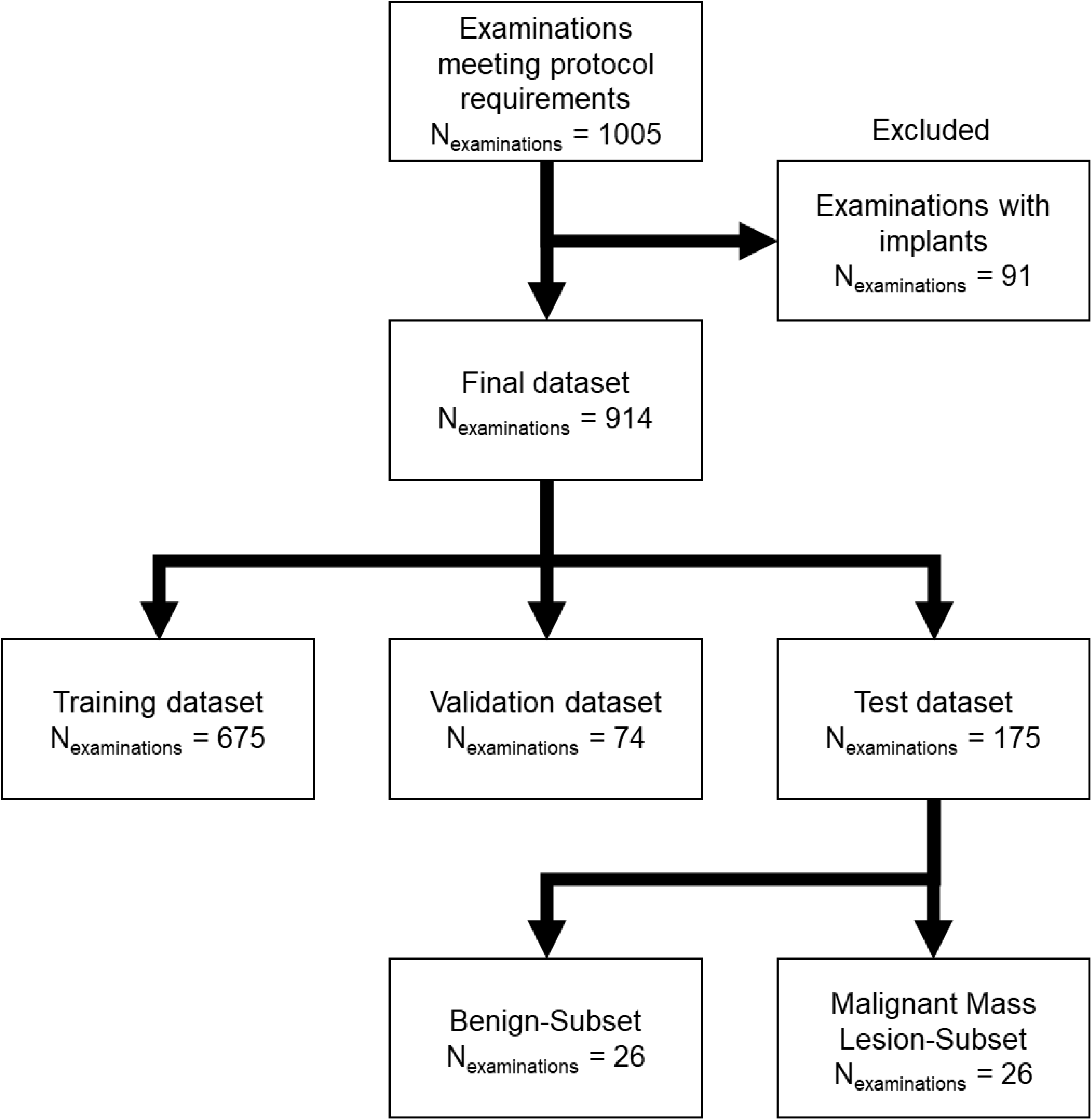
Study design flowchart. Protocol requirements for the examinations were defined as including uninterrupted acquisitions of: T1w, T1w-FS, T2w-FS, DWI with multiple b-values including 50,750 and 1500 s/mm^2^ as well as a 5 time-point T1w DCE acquisition.

### Data preprocessing

MRI acquisitions from each patient were extracted from the clinical PACS system and transferred to a local machine for preprocessing using Python (version 3.9.10) with SITK framework (version 2.2.1) as following. The Field of View (FoV) of the sequences was registered to smallest common volume among all of the sequences of the respective examination (in most cases this was the FoV provided by the DWI). Such registration ensures full correlation of voxel information among all acquisitions. Further all acquisitions were z-score normalized, clamped at values of [-1,15] and min-max rescaled, for the input images, to a domain [0,1] and for the T2w-FS acquisition to a domain of [-1,1]. Binary masks of the breast volume were generated using an in-house developed algorithm as presented in the supplement material.

### Neural network architecture and training

A 2D-U-Net network was implemented using Python with PyTorch, MONAI and Lightning frameworks as depicted in Figure 2 in similarity to [10].

**Figure 2:**
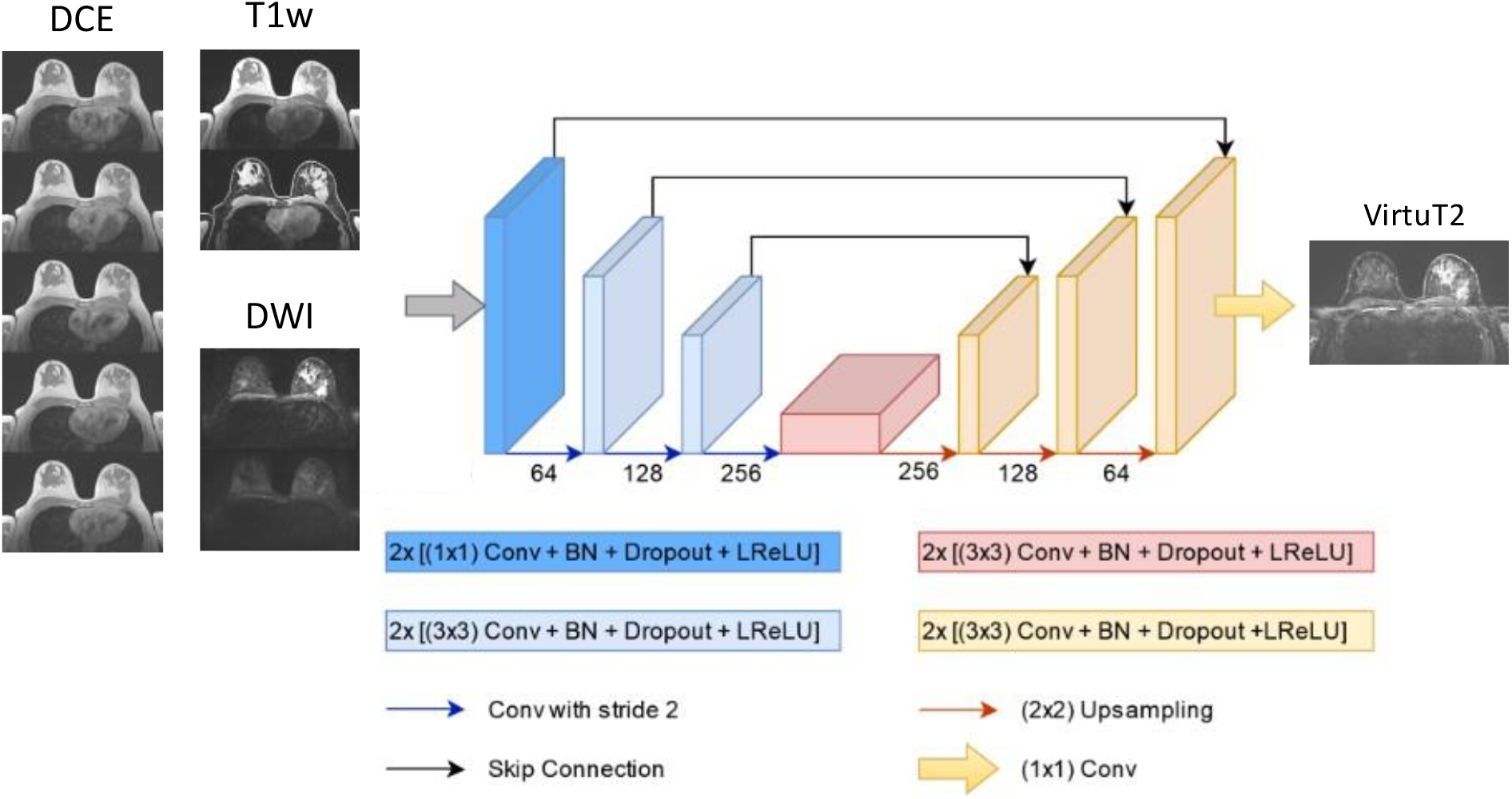
2D-U-Net architecture used for the generation of VirtuT2 images. The network comprises of 3 encoder- (in blue) and 3 decoder-stages (in orange) connected by a bottleneck stage (in red). The input of the network used during training consist of the T1w acquisition (both with and without fat-saturation), a series of T1w-contrast enhanced acquisition and of the DWI acquisitions with b-values of 50 and 750 s/mm^2^. The inputs of the network are passed in the initial stage through a 1×1 convolution layer. The initial encoder stage generates n=64 features and the feature number is multiplied by 2 after each encoder stage and the bottle-neck resulting in a n=512 maximal number of features. A skip connection between the respective levels of the encoder and decoder stages is introduced by concatenating the feature maps of the respective encoder-stage to the input of the decoder stage. Between the encoder-stages the down-sampling of the image dimensions is performed using a 2×2 convolution with stride of 2. The up-sampling between the decoder stages is performed using a transposed 2×2 convolution. The 2^nd^ and 3^rd^ encoder stage as well as the bottle-neck and the decoder stage consist of two sets of: 3×3 convolution layer followed by batch normalization, drop-out (with probability of 0.5) and leaky-rectified-linear-activation layers. The final encoder stage is followed by a 1×1 convolution layer and a tanh activation function.

During training, single slices of the T1w, T1w-FS, DWI acquisitions with b-values of 50 and 750 s/mm^2^ and all five T1w-CE acquisitions were used as input channels of the neural network. The corresponding slices of the T2w-FS acquisition were selected as the ground truth. Training was performed using a loss function which combines L1-norm with structural similarity measure (SSIM) in similarity to [17]:

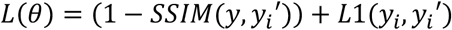

Training was performed on a dedicated work station using a single NIVIDIA V100 GPU (32GB RAM). For optimization an ADAM optimizer was used with a learning rate of 10^−3^. Training was performed for n=35 epochs using an n=32 batch-size.

#### Test Set Evaluation

After the training, virtual T2w-FS images were generated using the independent test dataset. The resulting images were then evaluated quantitatively and in a reader study, performed by two board-certified radiologists.

### Quantitative evaluation

Quantitative metrics evaluated within the full breast volume for the Virtu-T2 images were as follows: structural similarity index (SSIM) [18], peak signal-to-noise ratio (PSNR), histogram mutual information (HMI), normalized root mean square error (NRMSE), median symmetric accuracy (MEDSYMAC) and high frequency error norm (HFEN). All of the metrics were evaluated over the volume of the whole breast including breast muscle.

### Qualitative multi-reader study

To supplement the quantitative analysis a multi-reader study was performed on two subsets of the independent test set.

The two subsets were randomly selected from the full test set as following:

1. Subset A (n=26 examinations) comprising of breast MRI examinations with histopathologically confirmed malignant (mass) lesions.
2. Subset B (n=26 examinations) comprising of breast MRI examinations with benign findings only.

For both subsets a reader study was performed by two board certified radiologists (R1: D.H. 15 years of experience and R2: S.B. 10 years of experience in breast MRI). The readers performed two reading sessions for each subset with at least two weeks between each reading session. During each of the session readers were presented with a total of n=26 cases of multiparametric breast MRI of which n=13 cases each were presented including the VirtuT2 images or the original T2w-FS images. The information about the number of included Virtu-T2 and T2w-FS images in each reading session was withheld from the readers. The order in which cases were presented during both sessions was random. During the reading the following tasks were performed:

1. Readers evaluated whether the presented image overall provides T2w-FS image characteristics (e.g. fat saturated appearance of fat, hyperintense depiction of FGT proportions, hyperintense depiction of cysts) answering the question with a Yes/No.
2. Readers were asked to answer whether the presented image is an original T2w-FS acquisition or a VirtuT2 image.
3. Readers were asked to assess the diagnostic image quality (DIQ) using a 5-point Likert-like scale using the following scores: 5 = excellent (acceptable for diagnostic use), 4 = good (acceptable for diagnostic use), 3 = acceptable (acceptable for diagnostic use but with minor issues), 2 = poor (not acceptable for diagnostic use), or 1 = unacceptable (not acceptable for diagnostic use). For cases which were rated to have an unacceptable or poor diagnostic quality (scores 1 and 2) the readers were asked to indicate the reason for such rating as: artefact presence, image resolution, image blurring, lack of enhancement or other.

Additionally, for the first subset which included malignant mass lesions, in similarity to Kaiser score evaluation, the readers answered, if is a visible presence of edema around the largest lesion which could be associated with malignancy.

### Statistical Analysis

Statistical differences in the ordinal evaluations between the T2w-FS and VirtuT2 images were investigated using a Wilcoxon Signed Rank test for each of the readers, a p-value of 0.05 was considered significant. Inter-reader agreement between the two readers was evaluated using the Cohens’s kappa (ƙ) for the diagnostic image quality and for the presence of edema. Additionally, Intra-reader agreement between the T2w-FS and VirtuT2 images in regards to the presence of edema was also evaluated using Cohen’s kappa.

## RESULTS

### Patient cohort characteristics

Detailed information about the full patient cohort can be found in Table 2. Table 3 shows the frequency of BI-RADS adapted fibro-glandular tissue (FGT) and background parenchymal enhancement (BPE) classes in the independent test set. Among the n=26 cases included in the malignant subset used for reading, n=25 cases were identified as a breast cancer of no special type (NST) and n=1 as a mucinous carcinoma. Mean size of the malignant lesions was 20.5±17.1 mm (median 13.4 mm, n=8 cases with lesion size < 10 mm, n=7 cases with lesion size >20 mm).

**Table 2:** Demographics of the Training/Validation and Independent Test Cohorts.

|  |  |  |  |  |
| --- | --- | --- | --- | --- |
|  | Full dataset | Training/Validation set |  | Independent test set |
|  |  | Training | Validation |  |
| No. of patients | 816 | 567 | 74 | 175 |
| No. of examinations | 914 | 665 | 74 | 175 |
| Split ratio of examinations (%) | 100 | 72.7 | 8.1 | 19.1 |
| Age (year) | 52±12 | 50±12 | 52±12 | 51±14 |
| Routine BI-RADS score* |  |  |  |  |
| 0 | 20 | 14 | 2 | 4 |
| 1 | 3 | 2 | 0 | 1 |
| 2 | 440 | 328 | 36 | 76 |
| 3 | 70 | 53 | 3 | 14 |
| 4 | 106 | 68 | 10 | 28 |
| 5 | 64 | 50 | 3 | 11 |
| 6 | 211 | 150 | 20 | 41 |
| *The BI-RADS score refers to the highest BI-RADS score given for an examination during routine clinical reading; age is presented as means and standard deviations. |  |  |  |  |

**Table 3:** Fibro-glandular Tissue (FGT) and Background-Parenchymal Enhancement (BPE)

|  |  |  |  |
| --- | --- | --- | --- |
| FGT scores* |  | BPE scores* |  |
| Independent Test Set (n=175) |  |  |  |
| Almost entirely fat | 23<br>(13%) | Minimal | 62<br>(35%) |
| Scatter fibro-glandular tissue | 34<br>(19%) | Mild | 40<br>(22%) |
| Heterogeneous fibro-glandular tissue | 40<br>(22%) | Moderate | 32<br>(18%) |
| Extreme fibro-glandular tissue | 78<br>(44%) | Marked | 41<br>(23%) |
| *Scores given adapted to the ACR BI-RADS reporting recommendations |  |  |  |

### Evaluation of the VirtuT2 images

Table 4 shows the mean and standard deviation of the quantitative performance metrics for the VirtuT2 images in comparison to original T2w-FS.

**Table 4:** Quantitative Values for Comparison between VirtuT2 and T2w-FS images.

|  |  |
| --- | --- |
| metric | mean (± standard deviation) |
| SSIM ↑ | 0.87±0.02 |
| PSNR [dB] ↑ | 24.90±1.89 |
| HMI ↑ | 0.74±0.10 |
| NRSME [%] ↓ | 8.09±1.09 |
| MEDSYMAC [%] ↓ | 1.39±0.50 |
| HFEN ↓ | 0.87±0.07 |
| SSIM: structural similarity index; PSNR: peak signal-to-noise-ratio; NRMSE: normalized root mean square error; MEDSYMAC: median symmetrical accuracy; HFEN: high-frequency error norm; ↓ indicates lower values to suggest improvement of network performance; ↑ indicates higher values to suggest improvement of network performance; dB=decibel; | |

#### Identification of original and VirtuT2 images

All Virtu-T2 images (52/52) and T2w-FS images (52/52) were unanimously classified by both readers as providing typical characteristics of T2w-FS acquisitions (R1 52/52, R2 52/52). Yet, both readers were able to reliably identify the Virtu-T2 images with R1 correctly identifying 92.3% (48/52) and R2 correctly identifying 94.2% (49/52) of them. Accuracies for R1 in classifying original and VirtuT2 images was 96.2% (100/104) and 95.2% (99/104) for R2 respectively. Comparison of the T2w-FS and VirtuT2 images for both correctly and falsely identified cases by both readers are presented in Figure 3.

**Figure 3:**
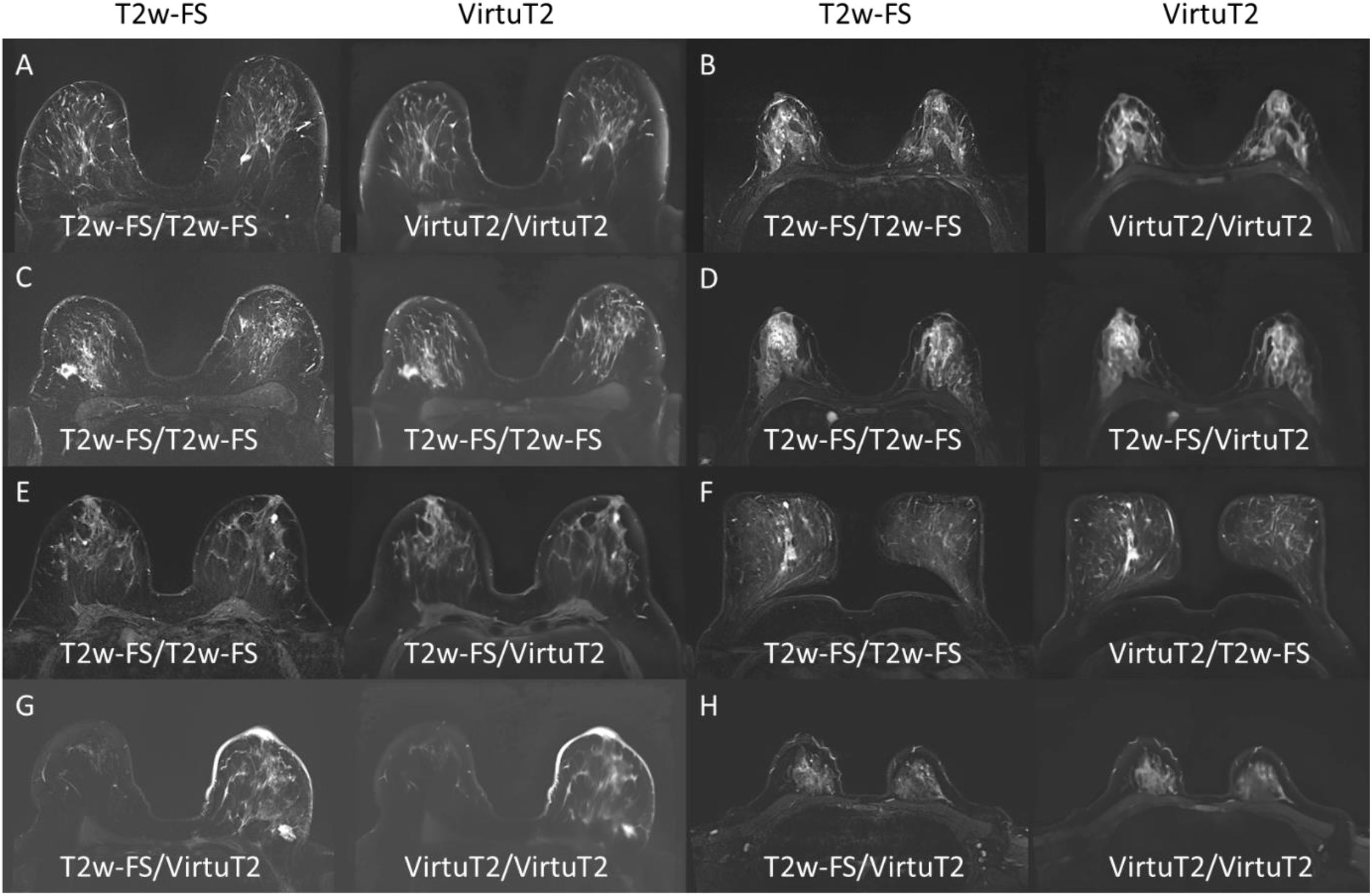
Example of images cases for which the two readers answered whether the respective image series looks like an original T2w-FS acquisition or a synthetic VirtuT2 image. The image names below each image indicate the respective readers answers with the R1 presented on the left side and R2 on the right side. The readers were able to identify the VirtuT2 images in most cases where significant blurring in the image is observed. In one case (C) both of the readers mistook a VirtuT2 image for a T2w-FS. Notably this image has a low level of blurring. In two cases (D and E) R1 mistook the VirtuT2 image for a T2w-FS. In two cases R2 (G and H) mistook an original T2w-FS image for a VirtuT2 image.

#### Image quality and diagnostic assessment

Inter-reader agreement for the evaluation of DIQ showed low agreement between the readers for T2w-FS images (ƙ=0.10) but a fair agreement for VirtuT2 images (ƙ=0.26) for the individual quality scores.

No significant difference in the DIQ scores given were found for R1 between the T2w-FS and VirtuT2 images (p=0.21). However, for R2 a significantly lower median could be observed for the VirtuT2 images (p<=0.001). Example of T2w-FS and VirtuT2 images with different DIQ scores are presented in Figure 4. Frequencies of the DIQ scores for both T2w-FS acquisition and for VirtuT2 are presented in Figure 5. For both of the lower end DIQ scores of 2 given by R2 the reason for the score was increased blurring.

**Figure 4:**
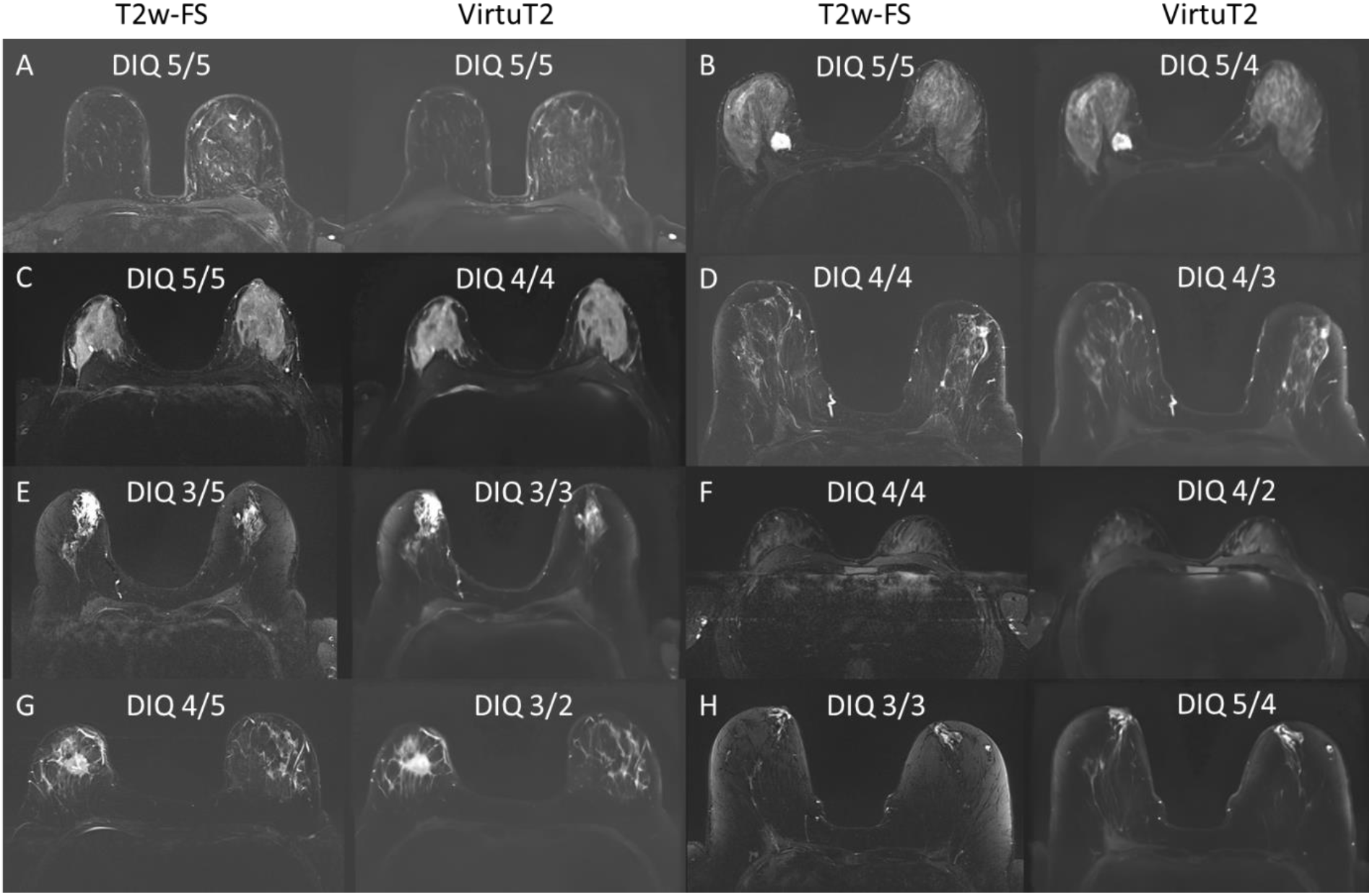
Example of images cases with different diagnostic image quality (DIQ) ratings given by the two readers. The scores above each image indicate the respective readers answers with the R1 presented on the left side and R2 on the right side. In most of the presented cases the DIQ was higher for the T2w-FS images then in the VirtuT2 images. An exception from this is image H in which the fat-supression didn’t work as intended and the DIQ of the T2w-FS was rated by both readers as only acceptable. At the same time two VirtuT2 images (F and G) were rated by R2 as having a poor (score 2) DIQ. No images in the whole cohort were rated as having am unacceptable quality.

**Figure 5:**
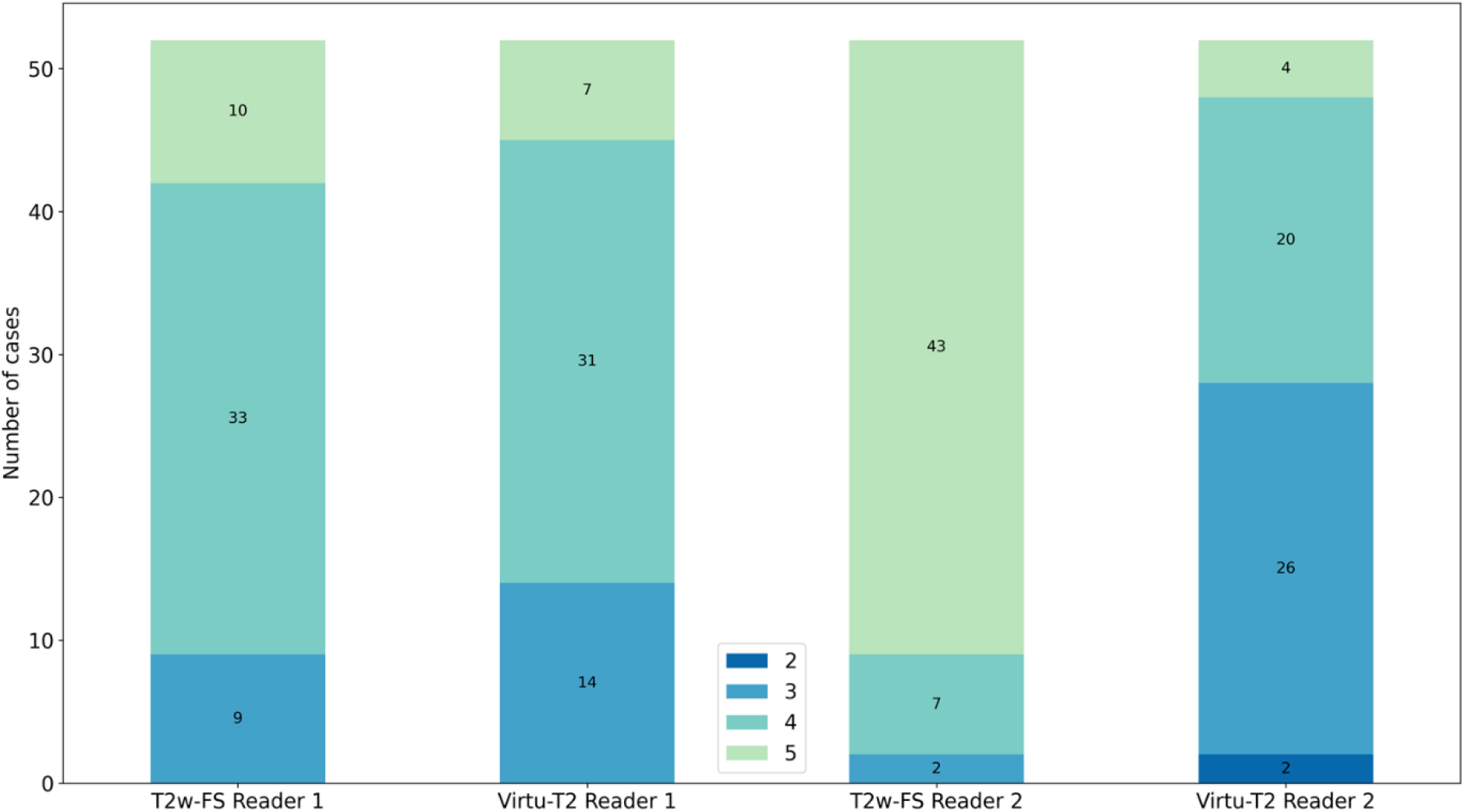
Frequencies of different DIQ scores given to the original T2w-FS and to the synthetic VirtuT2 images by the two readers. For Reader 1 no significant difference in the median of the score could be observed between the two methods (p=0.21). For Reader 2 a significantly larger portion of the T2w-FS acquisitions showed a higher DIQ then the VirtuT2 images (p<0.001).

#### Identification of presence of edema

With regard to the reading task of identification of presence of edema R1 identified it in n=12 cases and R2 in n=19 using the T2w-FS acquisitions. At the same time R1 identified the edema on n=12 cases on the VirtuT2 images and R2 on n=13. Moderate inter-rater agreement (ƙ=0.43) could be observed on the T2w-FS acquisitions. In comparison, for the VirtuT2 images only a fair agreement (ƙ=0.36) between the readers could be observed for this task. Between the T2w-FS and VirtuT2 images slight (ƙ=0.19) and fair (ƙ=0.23) intra-reader agreements could be observed for R1 and R2. If the presence of edema was identified on the original T2w-FS acquisition it could also be identified on the VirtuT2 images in only 50% (6/12) and 58% (11/19) of cases by R1 and R2 respectively. Figure 6 shows exemplary cases in which presence of edema was identified in either the original T2w-FS acquisitions or in the VirtuT2 image.

**Figure 6:**
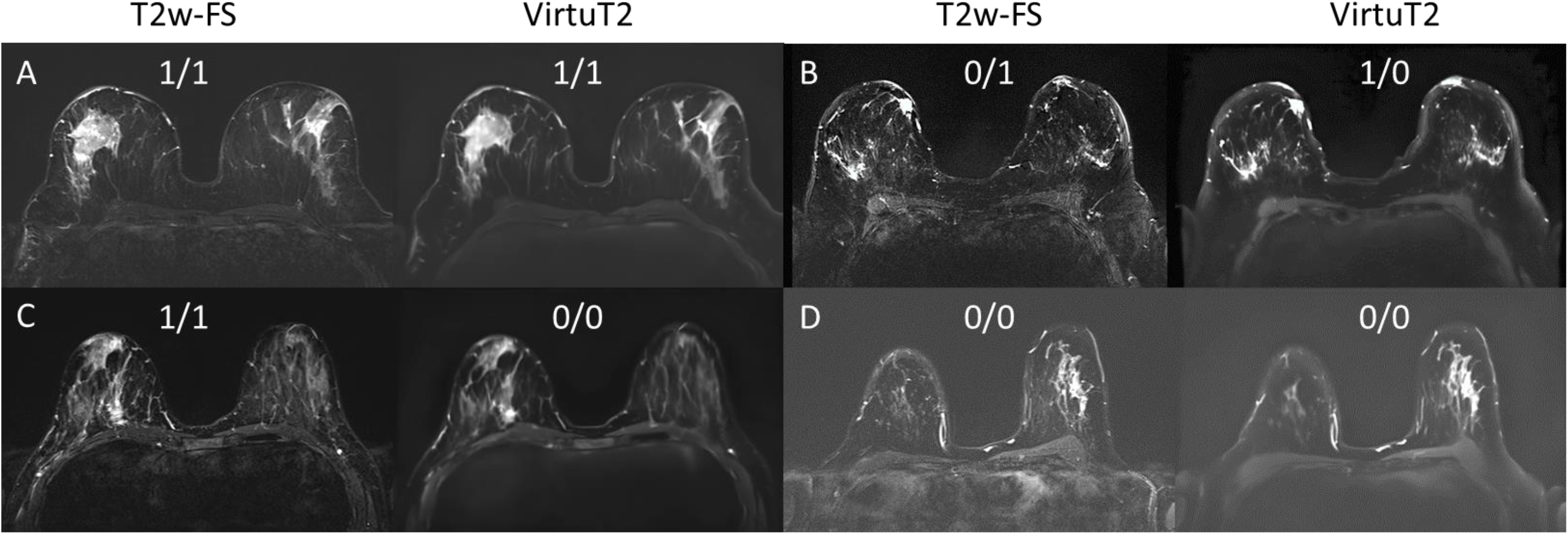
Example images of malignant cases with indications whether presence of edema was identified by R1/R2 respectively. A score of 1 indicates that the edema was present.

## DISCUSSION

This study demonstrates the generation of T2w-FS mimicking images from a multiparametric breast MRI protocol to be technically possible using a 2D-U-Net architecture. To our knowledge this study is the first in its kind showing that such application of deep learning is possible.

Our results indicated the VirtuT2 images to comprise typical T2w-FS image characteristics and high quantitative similarity metrics. However, the appearance of the VirtuT2 images differed enough from the original T2w-FS images for the readers to be reliably identified with a very high accuracy of 95-96%. Further, the reading revealed increased presence of blurring comprising diagnostic image quality in the VirtuT2 images.

The performance of the generated VirtuT2 images in regards to the quantitative similarity and error metrics were found in comparable ranges of virtually generated T1w contrast-enhanced images with an achieved SSIM of 0.87 and a PSNR of 24.90 [5-9]. Expanding the interpretability of the quantitative metrics we further included an evaluation of the HFEN, which is widely used to assess the level of image blurring [19].

Recent publications on breast MRI protocols indicated that T2w-FS acquisitions might take-up up to approximately 20% of the whole on-table examination time [2; 3]. Whilst only presenting a first feasibility of the VirtuT2 its development is motivated by its potential to further streamline scanner throughput times without compromising on the available image contrasts for diagnostic assessments.

However, it should be noted that the generated VirtuT2 images are not without flaws and the current feasibility status is still far away from clinical applicability, hindered amongst by the significant blurring of its generated images. The blurring affected the overall Diagnostic Image Quality (DIQ) scores for both of the two readers, although statistically significant difference could be observed only for R2. The high value of the HFEN of approx. 0.87 is well in agreement with the observed high level of blurring occurring in the VirtuT2 images. The blurring might be caused by the resolution of the DWI as relevant information source for the network. Another reason for the blurring might be the applied U-Net architecture, as encoder-decoder architectures are known for their blurry outputs if large variations in shape and scale is present in target regions [20; 21]. Such situation naturally occurs in T2w-FS images where large regions of fibro-glandular-tissue occur among areas speckled with small scale nodular or linear appearing tissue tracks. Interestingly the observed blurring appears in a similar fashion to blurring observed in the vCE example images in the works of Chung *et al*.[6]. This may point towards the architecture’s responsibility for the blurring as both this work and the work by Chung et al. are using encoder-decoder architectures. Nevertheless, for a definitive answer on this aspect further investigations of other architectures, for example through inclusion of attention mechanisms [7; 8] or a complete change of the architecture [7] will be necessary.

Further the VirtuT2 images represented visually visible edema only in 50%-58% of cases in which the edema was observed in the original T2w-FS acquisitions. However, this evaluation is strongly dependent on the reader as it can be seen from the only fair and moderate agreements between the readers as well as from the number of cases in which the edema was detected in the original acquisitions. Additionally due to its nature this evaluation does not have a reference value against which it could be compared against.

Final limitation is found in the relatively small dataset with only n=914 examinations performed on just two scanner types from a single manufacturer with both scanners using 3T magnetic field strength and a harmonized acquisition protocol with high-quality DWI acquisitions. These limitations affect the generalizability of our method and further investigations of this method on larger external datasets should be performed in future studies.

## CONCLUSION

Our study suggests that a neural network is able to technically generate images which mimic the contrast of T2w-FS acquisitions in breast MRI – however with increased blurring as compared to acquired T2w-FS data. Further research on this topic is necessary to overcome current limitations of this initial technical setup.

## Data Availability

Original image data used in this work are not publicly available to preserve individuals privacy under the European General Data Protection Regulation. The institution handling this data is the Institute of Radiology, University Hospital Erlangen.

## Abbreviations

CE: contrast enhanced
SSIM: structural similarity index
PSNR: peak signal-to-noise-ratio
HMI: histogram mutual information
NRMSE: normalized root mean square error
MEDSYMAC: median symmetrical accuracy
HFEN: high frequency error norm
DWI: diffusion weighted imaging

