## Supplement Material for "Feasibility to virtually generate T2 fat-saturated breast MRI by convolutional neural networks"

### Binary Mask generation

Binary masking was performed on the original T1w acquisition. Binary masks were calculated on multiple maximum intensity projections (MIP) in z-direction (slice direction). For each slice of the T1w data, an individual MIP was calculated. Those MIPs were binarized using a mean thresholding algorithm. To ensure homogeneous masks including the whole breast tissue, binary dilation with a disk-shaped kernel of 5 pixels in diameter followed by a binary closing operation was performed. To account for different anatomical properties in different slice locations of the volumetric data, MIPs were created using a selection of adjacent slices. The range of adjacent slices depending on the position in the volume is represented in Equation (1) below, where $n_{slices}$ define the range of slices considered for the MIP generation for a specific position. The masks were stored in NIfTI format with the spacing, direction and origin of the T1w data to enable a visualization with medical visualization applications.

$$n_{slices}(slice) = \begin{cases} [0,6] & if\ slice \leq 3 \\ [slice-3, slice+3] & if\ 3 < slice < 9 \\ [slice-3, slice+3] & if\ 87 < slice < 93 \\ [90,96] & if\ slice \geq 93 \\ [slice-8, slice+8] & otherwise \end{cases} \quad (1)$$

Supplement Figure 1 displays the results of breast volume masking on corresponding T1w slices for a representative case. The masks successfully include the whole breast tissue while the air around the patient is excluded as well as parts of the thorax, and most of the lung and heart tissue.

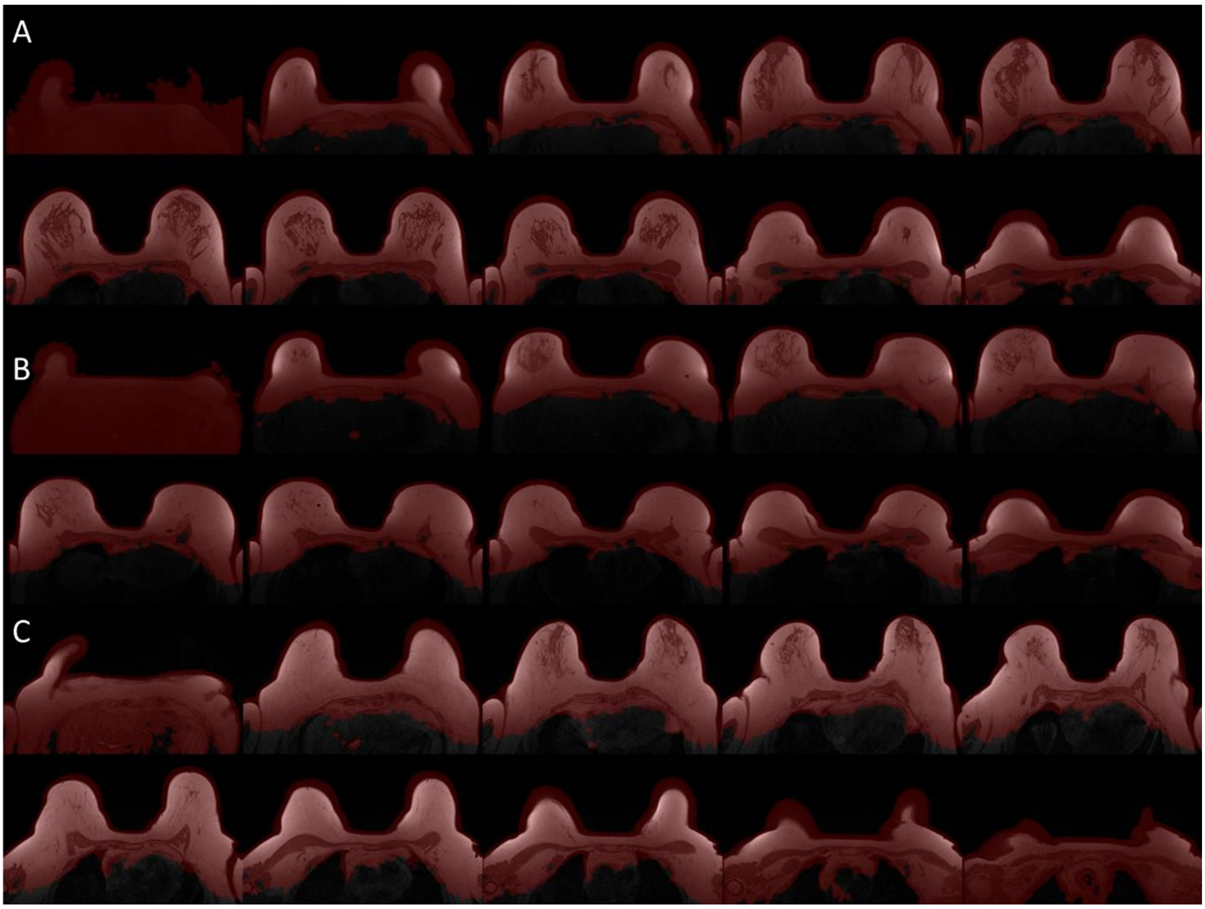


**Supplement Figure 1:** Example of binary masks generated using the in-house algorithm of every 10th slice for three examinations (A, B, C). It can be noted that the masks include both breast volume as well as parts of the heart, thorax and lung.
